# Evidence-Based, Cost-Effective Interventions To Suppress The COVID-19 Pandemic: A Systematic Review

**DOI:** 10.1101/2020.04.20.20054726

**Authors:** Carl-Etienne Juneau, Tomas Pueyo, Matt Bell, Genevieve Gee, Pablo Collazzo, Louise Potvin

## Abstract

**Background:** In an unparalleled global response, during the COVID-19 pandemic, 90 countries asked 3.9 billion people to stay home. Yet some countries avoided lockdowns and focused on other strategies, like contact tracing and case isolation. How effective and cost-effective are these strategies? We aimed to provide a comprehensive summary of the evidence on pandemic control, with a focus on cost-effectiveness.

**Methods:** Following PRISMA systematic review guidelines, MEDLINE (1946 to April week 2, 2020) and Embase (1974 to April 17, 2020) were searched using a range of terms related to pandemic control. Articles reporting on the effectiveness or cost-effectiveness of at least one intervention were included and grouped into higher-quality evidence (randomized trials) and lower-quality evidence (other study designs).

**Results:** We found 1,653 papers; 62 were included. Higher-quality evidence was only available to support the effectiveness of hand washing and face masks. Modelling studies indicated that these measures are highly cost-effective. For other interventions, lower-quality evidence suggested that: (1) the most cost-effective interventions are swift contact tracing and case isolation, surveillance networks, protective equipment for healthcare workers, and early vaccination (when available); (2) home quarantines and stockpiling antivirals are less cost-effective; (3) social distancing measures like workplace and school closures are effective but costly, making them the least cost-effective options; (4) combinations are more cost-effective than single interventions; (5) interventions are more cost-effective when adopted early and for severe viruses like SARS-CoV-2. For H1N1 influenza, contact tracing was estimated to be 4,363 times more cost-effective than school closures ($2,260 vs. $9,860,000 per death prevented).

**Conclusions:** A cautious interpretation of the evidence suggests that for COVID-19: (1) social distancing is effective but costly, especially when adopted late and (2) adopting as early as possible a combination of interventions that includes hand washing, face masks, ample protective equipment for healthcare workers, and swift contact tracing and case isolation is likely to be the most cost-effective strategy.

**Funding:** LP holds the Canada Research Chair in Community Approaches and Health Inequalities (CRC 950-232541). This funding source had no role in the design, conduct, or reporting of the study.

## INTRODUCTION

On March 11, 2020, the World Health Organization (WHO) characterized COVID-19 as a pandemic. The virus then infected exponentially more men and women worldwide. In an unparalleled global response, more than 90 countries or territories have asked about half of the world’s population to stay home (Sandford, 2020). During that time, over 1.5 billion (almost 90%) of the world’s students were affected by nationwide school closures (WHO, 2020b). Other countries focused on other interventions, such as contact tracing and case isolation. These have been estimated to be 4,363 times more cost-effective than school closures for H1N1 influenza ($2,260 vs. $9,860,000 per death prevented) (Madhav et al. 2017). Indeed, closing school is costly—$10 to $47 billion for 4 weeks in the US alone (Lempel et al. 2009). As countries around the world are faced with the challenge of balancing public health interventions with economic, ethical, social, and legal considerations, evidence on the effectiveness and cost-effectiveness of these interventions is urgently needed to guide policy and avoid unnecessary harm.

An earlier systematic review of non-pharmaceutical interventions to reduce influenza transmission in adults included only randomized trials, analyzed 7 studies, and concluded that the evidence was lacking for most interventions (Smith et al. 2015). While we do not dispute this conclusion when looking only at randomized trials, we would argue that as urgent decisions of unknown cost-effectiveness are made in reaction to the COVID-19 pandemic, some evidence, even of lower quality, is better than no evidence at all. Therefore, we included a broad range of study designs in this review, and aimed to provide a comprehensive summary of the evidence on epidemic control, with a focus on cost-effectiveness.

## METHODS

We report this systematic review according to PRISMA guidelines (Moher et al. 2009). Reviews (all types), randomized trials, observational studies, and modelling studies were included. Articles reporting on the effectiveness or cost-effectiveness of at least one intervention were included. We defined effectiveness as success in producing the desired outcome, and cost-effectiveness as doing so with minimum economic cost (in dollar value). Articles in English, French, Spanish, and Portuguese were included. Studies of sexually transmitted infections (e.g. syphilis) and mosquito-borne diseases (e.g. dengue) were not included. Abstracts, case reports, and conferences proceedings were also excluded. Hand washing and face masks were the focus of a number of reviews (Jefferson et al. 2011; Smith et al. 2015; MacIntyre et al. 2015) and a recent meta-analysis (Saunders-Hastings et al. 2017), so individual studies of their effectiveness were also excluded. Likewise for school closures (Jackson et al. 2014; Rashid et al. 2015; Bin Nafisah et al. 2018; Viner et al. 2020). MEDLINE (1946 to April week 2, 2020) and Embase (1974 to April 17, 2020) were searched using the terms “non-pharmaceutical interventions”, “outbreak control”, “outbreak interventions”, “epidemic control”, “epidemic interventions”, “pandemic control”, and “pandemic interventions” (last search: April 19, 2020). Reference lists and PubMed related articles of included studies were reviewed to find additional articles. Titles were screened by a single investigator. Abstracts and full texts were screened by two investigators. Discrepancies were solved by mutual agreement. Key methodological characteristics and findings of studies were recorded in a spreadsheet. These included first author, date of publication, study design, interventions, and key findings. Quality assessment was limited to grouping studies based on design into two categories: higher quality (randomized trials) and lower quality (other designs). Meta-analysis was not feasible due to the heterogeneous set of interventions studied, as well as substantial differences in study designs, outcomes, and effect measures. We described results narratively.

## RESULTS

### Result of the search

A total of 2,742 papers were found. Removing duplicates left 1,653. We retained 622 based on title, 137 based on abstract, and 39 based on full text. We found 23 additional studies via reference lists and PubMed related articles searches (eFigure in the Supplement). Therefore, a total of 62 studies were included (Table 1). Randomized trial evidence was only available to support the effectiveness of hand washing and face masks (Jefferson et al. 2011; Smith et al. 2015; MacIntyre et al. 2015; Saunders-Hastings et al. 2017). For other interventions, only lower-quality (observational and modelling) evidence was available.

### Cost-effectiveness of interventions

Pasquini-Descomps et al. (2017) conducted a systematic review of the cost-effectiveness of interventions in H1N1 influenza. They found 18 studies covering 12 interventions: disease surveillance networks (very cost-effective), contact tracing and case isolation (very cost-effective), face masks (very cost-effective), preventive measures in hospitals (cost-effective), antiviral treatment (cost-effective), antiviral prophylaxis (cost-effective), low efficiency vaccination (cost-effective if timed before cases peak), high efficiency vaccination (cost-effective if timed before cases peak), stockpiling antiviral medicine (cost-effective for high-income countries), quarantining confirmed cases at home (cost-effective for viruses with a case fatality rate of 1%, not cost-effective for viruses with a case fatality rate of 0.25%), self-isolation at home (cost-effective with a case fatality rate of 1%, not cost-effective with a case fatality rate of 0.25%), and school closure (not cost-effective). Based on these findings, Madhav et al. (2017) estimated that for H1N1 influenza, contact tracing was 4,363 times more cost-effective than school closures ($2,260 vs. $9,860,000 per death prevented). Other systematic reviews found that school closures did not help control of the 2003 SARS epidemic in China, Hong Kong, and Singapore and would prevent only 2-4% of COVID-19 deaths (Viner et al. 2020); reduced the peak of epidemics by 29.65% on average and were more effective when timed early (Bin Nafisah et al. 2018); are most effective when they cause large reductions in contact, when the basic reproduction number is below 2, and when attack rates are higher in children than in adults (Jackson et al. 2014); and appeared to be moderately effective in reducing the transmission of influenza and in delaying the peak of an epidemic, but were associated with very high costs (Rashid et al. 2015). Differences in publication date, virus transmissibility, and study selection may explain the discrepancies among these reviews.

Using data from Wang et al. (2012), Pasquini-Descomps et al. (2017) found that contact tracing and case isolation was one of the most cost-effective interventions to control H1N1 in Hubei, China (less than $1,000 per disability-adjusted life year). In a simulation study, Hellewell et al. (2020) found that in most scenarios, highly effective contact tracing and case isolation would be enough to control a new outbreak of COVID-19 within 3 months. Transmissibility was an important factor: when Ro = 2.5, 80% of contacts needed to be traced and isolated. Timing was another important factor: with five initial cases, there was a greater than 50% chance of achieving control, even at lower contact-tracing levels. However, at 40 initial cases, control was much less likely. Similarly, any delay from symptom onset to isolation decreased the probability of control, highlighting the need for swift action. Further, Armbruster and Brandeau (2007) found that contact tracing is cost-effective only when population prevalence is still low (e.g. under 8% for tuberculosis). In a systematic review, Halton et al. (2013) found that contact tracing and progressively earlier isolation of probable SARS cases were associated with control of SARS outbreak in Southeast Asia. In another review, Bell et al. (2006) recommended contact tracing and case isolation at the start of an outbreak, but not in the late phase, when there is “increased and sustained transmission in the general population”. In a modelling study, Zhang et al. (2015) found that voluntary self-isolation at symptom onset can achieve the same level of effectiveness as antiviral prophylaxis, but that this strategy had a limited effect on reducing transmission when delayed by two days. Young et al. (2019) also found that delays could prevent case isolation from stopping incipient outbreaks. Li et al. (2013) found that in 2009 H1N1, quarantine of close contacts in Beijing reduced confirmed cases by a factor of 5.6. However, since H1N1 was mild, they concluded that this was not an economically effective measure. In another modelling study, Tuncer et al. (2018) found that social distancing had the most impact on the 2014 Ebola epidemic in Liberia, followed by isolation and quarantine. Case isolation, household quarantine, and contact tracing were the most effective interventions in four other modelling studies (Becker et al. 2005; Sang et al. 2012; Chen et al. 2018; MacIntyre et al. 2019). Collectively, in the context of COVID-19, these studies suggest that these interventions can be effective and cost-effective, and highly so when implemented early and executed swiftly.

Saunders-Hastings et al. (2017) carried out a systematic review and meta-analysis of personal protective measures to reduce pandemic influenza transmission. Meta-analyses suggested that regular hand hygiene provided a significant protective effect (OR = 0.62; 95% CI 0.52–0.73). Face masks had a non-significant protective effect (OR = 0.53; 95% CI 0.16–1.71) which became significant (OR = 0.41; 95% CI 0.18–0.92) when randomized control trials and cohort studies were pooled with case–control studies (this also decreased heterogeneity). In an earlier systematic review, Jefferson et al. (2011) also found a protective effect of masks. Overall, they were the best performing intervention across populations, settings, and threats. Similarly, in a narrative review, MacIntyre et al. (2015) drew on evidence from randomized community trials to conclude that face masks do provide protection against infection in various community settings, subject to compliance and early use. Differences in publication date, search strategy, and study selection criteria may explain the discrepancies among these reviews. Tracht et al. (2012) estimated savings of $573 billion if 50% of the US population used masks in an unmitigated H1N1 epidemic. For hand washing, Townsend et al. (2017) estimated that a national behaviour change program in India would net $5.6 billion (3.4-8.6), a 92-fold return on investment. A similar program in China would net $2.64 billion (2.08-5.57), a 35-fold return on investment.

Preventive measures in hospitals include use of personal protective equipment for healthcare workers in direct contact with suspected patients. Dan et al. (2009) estimated that this measure was cost-effective for H1N1 ($23,600 per death prevented). However, adopting a wider set of measures (full personal protective equipment, restricting visitors, and cancelling elective procedures) was much less cost-effective ($2,500,000 per death prevented). Similarly, Lee et al. (2011) found that increasing hand hygiene, use of protective apparel, and disinfection are the most cost-saving interventions to control a hospital outbreak of norovirus. If they are not adequately protected, healthcare workers can contribute disproportionately to the transmission of the infection (Barnes et al. 2007).

Suphanchaimat et al. (2020) found that influenza vaccination for prisoners in Thailand was cost-effective. The incremental cost-effectiveness ratio of vaccination (compared with routine outbreak control) was $1282 to $1990 per disability-adjusted life year. Shiell et al. (1998) also found that vaccination (for measles) was cost-effective ($32.90 marginal cost per case prevented). Prosser et al. (2011) also found that H1N1 vaccination in the US was cost-effective under many assumptions if initiated prior to the outbreak. Incremental cost-effectiveness ratios ranged from $8,000 to $52,000 per quality-adjusted life year for persons aged 6 months to 64 years without high-risk conditions. The authors noted that all doses (two for some children, one for adults) should be delivered before the epidemic peak. Similarly, in a modelling study, Nguyen et al. (2018) found that vaccination should be administered five months before to one week after the start of an epidemic to be cost-effective. If vaccine supplies are limited, Lee et al. (2010) found that priority should be given to at-risk individuals, and to children within high-risk groups. Likewise, Van Genugten et al. (2003) estimated similar results from vaccinating the entire population vs. only at-risk groups. Herrera-Diestra and Meyers (2019) found that vaccinating based on the number of infected acquaintances is expected to prevent the most infections while requiring the fewest intervention resources. Optimal control modelling studies also suggest that early intervention and vaccination are more cost-effective, and that interventions before vaccines are available need to be balanced with the potential gains of future vaccines or the potential for multiple outbreaks (Handel et al. 2006; Lin et al. 2010; Buonomo and Messina, 2012).

In another systematic review of economic evaluations, Pérez Velasco et al. (2012) examined 44 studies and found that combinations of pharmaceutical and non-pharmaceutical interventions were more cost-effective than vaccines and/or antivirals alone. Reducing non-essential contacts, using pharmaceutical prophylaxis, and closing schools was the most cost-effective combination for all countries. However, quarantine for household contacts was not cost-effective, even in low- and middle-income countries. A modelling study by Day et al. (2006) suggested that quarantine (of all individuals who have had contact with an infected individual) would be beneficial only when case isolation is ineffective, when there is significant asymptomatic transmission, and when the asymptomatic period is neither very long, nor very short.

Perlroth et al. (2010) estimated the health outcomes and costs of combinations of 4 social distancing strategies and 2 antiviral medication strategies. For a virus with a case fatality rate of 1% and a reproduction number of 2.1 or greater, school closure alone was the least cost-effective intervention and cost $32,100 per case averted. Antiviral treatment ($18,200), quarantine of infected individuals ($15,300), and adult and child social distancing ($5,600) had increasing levels of cost-effectiveness. However, combining interventions was more cost-effective, and the most cost-effective combination included adult and child social distancing, school closure, and antiviral treatment and prophylaxis ($2,700 per case). However, the same combination without school closure was more cost-effective for milder viruses (case fatality rate below 1%, reproduction number 1.6 or lower). If antivirals are not available, the combination of adult and child social distancing and school closure was most effective. Similarly, in another modelling study, Bolton et al. (2012) found that a combination of non-pharmaceutical interventions proved as effective as the targeted use of antivirals.

In a similar study of cost-effectiveness, Saunders-Hastings et al. (2017b) examined a range of interventions (school closure, community-contract reduction, hand hygiene, face mask, voluntary isolation, quarantine, vaccination, antiviral prophylaxis, antiviral treatment) in response to a simulated pandemic similar to 1957 H2N2. In a population of 1.2 million, with no intervention, 9,421 life-years were lost.. Vaccination plus antiviral treatment was the most cost-effective intervention (cost per life-year saved: $2,581). However, it still led to 3,026 life-years lost. Only 1,607 life-years were lost at a marginally higher cost ($6,752 per life-year) with a combination of interventions including community-contact reduction, hand hygiene, face masks, voluntary isolation, and antiviral therapy. Combining all interventions saved the most lives (only 267 life-years lost), but was very costly ($199,888 per life-year saved) due to school closure and work days lost.

Halder et al. (2011) aimed to determine the most cost-effective interventions for a pandemic similar to H1N1. They found that a combination of interventions was most cost-effective. This combination included treatment and household prophylaxis using antiviral drugs and limited duration school closure ($632 to $777 per case prevented). If antiviral drugs are not available, limited duration school closure was significantly more cost-effective compared to continuous school closure. Other social distancing strategies, such as reduced workplace attendance, were found to be costly due to productivity losses. Closing school for 2 to 4 weeks without other interventions did not cost much more than doing nothing but gave a significant 34% to 37% reduction in cases if optimally timed.

### Studies on intervention effectiveness without cost-effectiveness analysis

Smith et al. (2015) carried out a systematic review of non-pharmaceutical interventions to reduce the transmission of influenza in adults. Only randomized trials were included and 7 studies met all selection criteria. The authors found that positive significant interventions included professional oral hygiene intervention in the elderly and hand washing, and noted that home quarantine may be useful, but required further assessment.

Jefferson et al. (2011) conducted a Cochrane systematic review of physical interventions to interrupt or reduce the spread of respiratory viruses. They found that the highest quality randomized cluster trials suggested this could be achieved by hygienic measures such as handwashing, especially around younger children. They recommended that the following effective interventions be implemented, preferably in a combined fashion, to reduce transmission of viral respiratory disease: frequent handwashing with or without adjunct antiseptics; barrier measures such as gloves, gowns and masks with filtration apparatus; and suspicion diagnosis with isolation of likely cases.

Lee et al. (2009) carried out a systematic review of modelling studies quantifying the effectiveness of strategies for pandemic influenza response. They found that combinations of strategies increased the effectiveness of individual strategies and could reduce their potential negative impact. Combinations delayed spread, reduced overall number of cases, and delayed and reduced peak attack rate more than individual strategies. Similar results were found by Martinez and Das (2014). In another systematic review of 12 modelling and three epidemiological studies, Ahmed et al. (2018) found that workplace social distancing reduced cumulative influenza attack rate by 23%. It also delayed and reduced peak attack rate.

Pan et al. (2020) examined associations between public health interventions and the epidemiology of COVID-19 in Wuhan, China. Traffic restrictions, cancellation of social gatherings, and home quarantines were associated with reduced transmission, but were not sufficient to prevent increases in confirmed cases. These were reduced and estimates of the effective reproduction number fell below 1 only when additional interventions were implemented. Those included hospital-based measures (designated hospitals and wards, use of personal protective equipment, increased testing capacity, accelerated reporting, and timely medical treatment) and community-based interventions (quarantine of presumptive cases and quarantine of confirmed cases of their close contacts in designated facilities).

Markel et al. (2007) examined non-pharmaceutical interventions in US cities during the 1918-1919 influenza pandemic (isolation or quarantine, school closure, public gathering ban). They found that all 43 cities in the study adopted at least one of these interventions, and that 15 cities applied all three. The most common combination (school closure and public gathering bans) was implemented in 34 cities (79%) for a median duration of 4 weeks and was significantly associated with reductions in weekly excess death rate. Cities that implemented interventions earlier had greater delays in reaching peak mortality (Spearman r=-0.74, P<0.001), lower peak mortality rates (Spearman r=0.31, P=.02), and lower total mortality (Spearman r=0.37, P=.008). There was a significant association between increased duration of interventions and a reduced total mortality burden (Spearman r=-0.39, P=.005). Another similar, historical study of US cities found that early intervention was associated with lower mortality (R^2^=0.69, P<0.01) (Bootsma and Ferguson, 2007).

Ishola and Phin (2011) reviewed the literature on mass gatherings. They found 24 studies and cautiously concluded that there is some evidence to indicate that mass gatherings may be associated with an increased risk of influenza transmission. In a more recent systematic review, Rainey et al. (2016) found that mass gathering-related respiratory disease outbreaks were relatively rare between 2005 and 2014 in the US. They concluded that this could suggest— perhaps surprisingly—low transmission at most types of gatherings, even during pandemics. Similarly, in a US survey of 50 state health departments and 31 large local health departments, Figueroa et al. (2017) found that outbreaks at mass gatherings were uncommon, even during the 2009 H1N1 pandemic. In a modelling study, Shi et al. (2010) found that mass gatherings that occur within 10 days before the epidemic peak can result in a 10% relative increase in peak prevalence and total attack rate. Conversely, they found that mass gatherings may have little effect when occurring more than 40 days earlier or 20 days after the infection peak (when initial Ro = 1.5). Thus the timing of mass gatherings might explain the apparent lack of evidence in support of their ban.

Recently, Zhao et al. (2020) quantified the association between domestic travel out of Wuhan, China, and the spread of SARS-CoV-2. Using location-based data, they estimated that each increase of 100 in daily new cases and daily passengers departing from Wuhan was associated with an increase of 16.25% (95% CI: 14.86–17.66%) in daily new cases outside of Wuhan. Ryu et al. (2020) conducted a systematic review of international travel restrictions, screening of travelers, and border closure. They examined 15 studies and concluded that the evidence did not support entry screening as an effective measure, and that travel restrictions and border closures would have limited effectiveness in controlling pandemic influenza. In another systematic review, Mateus et al. (2014) concluded that the evidence did not support travel restrictions as an isolated intervention for the containment of influenza, and that restrictions would be extremely limited in containing the emergence of a pandemic virus. Chong and Ying Zee (2012) modelled the impact of travel restrictions on the 2009 H1N1 pandemic in Hong Kong. They estimated that restricting air travel from infected regions by 99% would have delayed the epidemic peak by up to two weeks. Restricting both air and land travel (from China) delayed the peak by about 3.5 weeks. However, neither 90% nor 99% travel restrictions reduced the epidemic magnitude by more than 10%, and antiviral treatment and hospitalization of infectious subjects were found to be more effective than travel restrictions. Chinazzi et al. (2020) modelled the impact of travel limitations on the spread of COVID-19. They estimated that the travel quarantine of Wuhan delayed the overall epidemic progression by 3 to 5 days in mainland China, and reduced international case importations by nearly 80% until mid-February. In addition, sustained 90% travel restrictions to and from China only modestly affected the epidemic trajectory, unless combined with a 50% or higher reduction of transmission in the community. Bell et al. (2006b) point out that screening international travelers who depart infected countries (instead of all travelers entering all countries) would be a better use of resources. Case in point: Zhang et al. (2011) reported that in 2009 H1N1, only 132 of the 600,000 travelers who underwent border entry screening in Beijing were infected (0.02%). Travel limitations may be more effective when neighbouring countries fail to implement adequate outbreak control efforts (Caley et al. 2007; Bwire et al. 2016).

We found little evidence to support the following interventions: (1) communicating health risk and promoting disease control measures in low and middle-income countries (evidence not conclusive according to a review by Schiavo et al. 2014); (2) screening to contain spread, at the borders or locally (even under best-case assumptions, more than half of infected people would be missed, according to a modelling study by Gostic et al. 2020).

## DISCUSSION

This systematic review aimed to provide a comprehensive summary of the evidence on pandemic control, with a focus on cost-effective interventions in the context of COVID-19. Randomized trial evidence was only available to support the effectiveness of hand washing and face masks. Modelling studies further suggested that these measures are highly cost-effective. For other interventions, only evidence from observational and modelling studies was available. This lower-quality evidence suggests that overall, when timed appropriately, the following interventions are likely to be highly cost-effective: contact tracing and case isolation, protective equipment for healthcare workers, and vaccination prior to the outbreak (when available). Surveillance networks and protective equipment for healthcare workers also appear to be cost-effective. Home quarantine for confirmed cases and stockpiling antivirals appear less cost-effective. The least cost-effective interventions appear to be social distancing measures like workplace and school closures. The evidence suggests that these are more cost-effective for severe viruses like SARS-CoV-2, and when timed early in the outbreak. Vaccination past the peak of infections and longterm school closures late in the outbreak appear less cost-effective, underscoring the importance of timing.

As many authors have noted, the cost-effectiveness of interventions depends on virus severity. For SARS-CoV-2, estimates of case fatality rate range from 1% to 7.2% (Onder et al. 2020), making it rather severe. The cost-effectiveness of interventions also depends on their timing. Taking this into account, we propose a 3-stage framework for pandemic control interventions (Figure 1). Interventions are shown from top (most cost-effective) to bottom (least cost-effective), according to the three stages described by Madhav et al. (2017) as prepandemic, spark, and spread (shown from left to right). A complete description is found in the Supplement.

**Figure 1.**
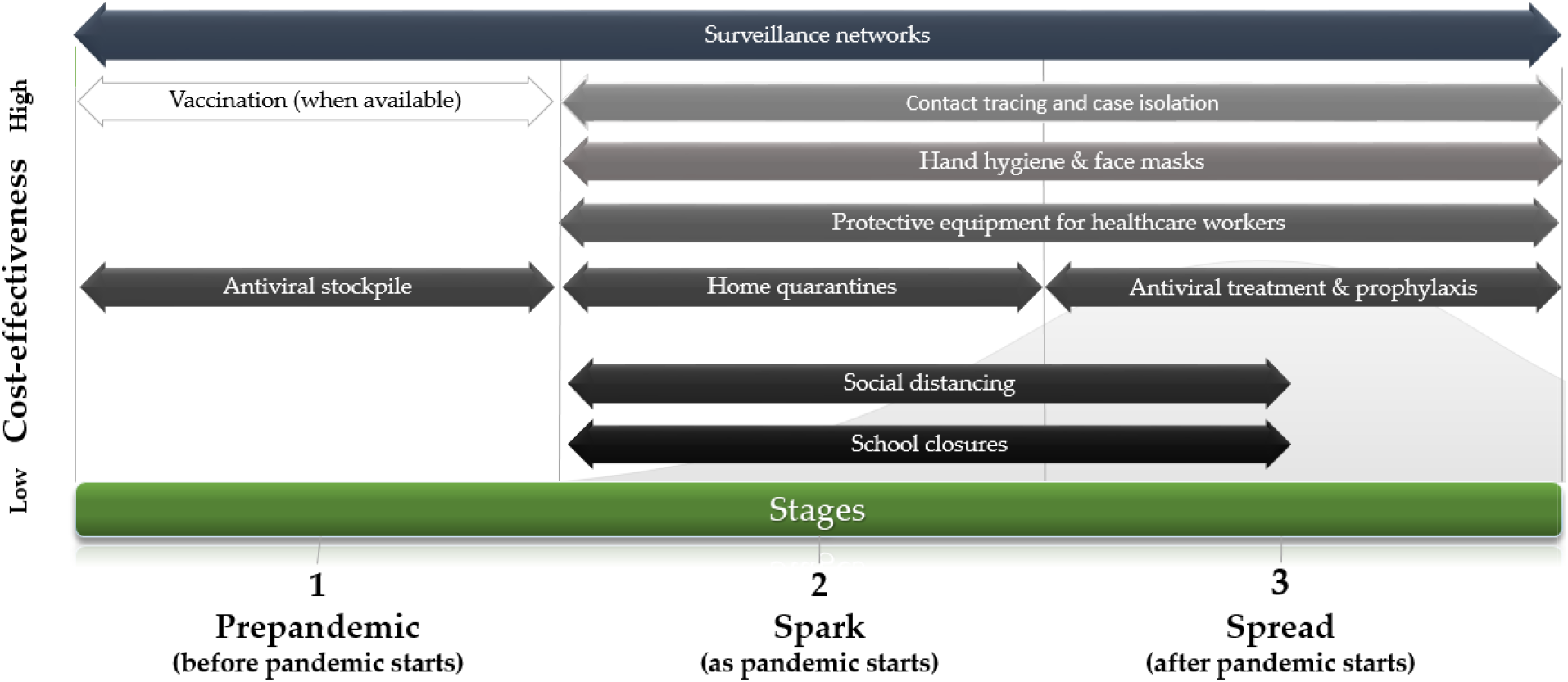
Cost-effectiveness of interventions in COVID-19, by stage In Figure 1, we propose a 3-stage framework for cost-effective pandemic control. Interventions are shown from top (most cost-effective) to bottom (least cost-effective), according to the stages described by Madhav et al. (2017) as prepandemic, spark, and spread (shown from left to right). According to this framework, surveillance networks are highly cost-effective, should be established before the pandemic starts (stage 1), and maintained through stages 2 and 3. Vaccination, when available, should occur before the pandemic, or as early as possible. Antivirals can be stockpiled cost-effectively in high-income countries. As the pandemic starts (stage 2), early contact tracing and case isolation is the most cost-effective intervention. It may be sufficient to contain the outbreak. If the outbreak is not contained, hand hygiene, face masks, and protective equipment for healthcare workers are all highly cost-effective. If these measures are not sufficient, home quarantines, social distancing, and school closures are all effective, albeit increasingly costly measures. Assuming a 1 to 2% case fatality rate for COVID-19, these measures are likely to be cost-effective nonetheless, especially if implemented early. As COVID-19 spreads (stage 3), and especially past the peak, the costliest interventions can be replaced cost-effectively by a combination of interventions centered on swift contact tracing and case isolation. Once antivirals are available, they can also replace the costlier interventions cost-effectively.

### Strengths and limitations

This review has one key strength: it included a broad range of study designs to provide a comprehensive summary of the evidence. This could however also be viewed as a limitation, as it includes evidence of both high and low quality. Lower-quality evidence should be interpreted with caution. Still, as randomized trial evidence was not available for most epidemic control interventions, and as COVID-19 forces urgent decision making, we submit that some evidence, even of lower quality, is better than no evidence at all. In addition, this review has a number of limitations. First, because of time constraints, our search was limited to two databases (MEDLINE and Embase) and we did not examine risk of bias. Second, most studies focused on H1N1 and other viruses, not SARS-CoV-2. Third, estimates of COVID-19 case fatality rate are subject to substantial uncertainties. They are likely to change as more data emerge. Should the true rate be high, all interventions would be more cost-effective. Conversely, should it be low, costly interventions such as school closures may not be cost-effective at all.

## Conclusions

Hand washing and face masks were the only measures supported by higher-quality evidence. Other interventions were supported by lower-quality evidence. In the context of COVID-19, a cautious interpretation suggests that (1) social distancing is effective but costly, especially when adopted late and (2) adopting as early as possible a combination of interventions that includes hand washing, face masks, swift contact tracing and case isolation, and protective equipment for healthcare workers is likely to be the most cost-effective strategy.

## Data Availability

Data extracted from cited work.

## Funding

LP holds the Canada Research Chair in Community Approaches and Health Inequalities (CRC 950-232541). This funding source had no role in the design, conduct, or reporting of the study.

## Conflicts of interests

CEJ, TP, MB, PC, and LP declare no conflict of interest. GG holds a contractual position with the Millar Group (a provider of personal protective equipment) and executive roles at Panacea Health Solutions and Angular Momentum (providers of diabetes and corporate wellness programs).

## Transparency declaration

The corresponding author (CEJ) affirms that this manuscript is an honest, accurate, and transparent account of the study being reported; that no important aspects of the study have been omitted; and that any discrepancies from the study as planned have been explained.

## Authors’ contributions

CEJ, TP, and LP designed the study. CEJ, GG, MB, and PC searched and analyzed the literature. CEJ and TP interpreted the findings. CEJ wrote the first draft. All authors revised drafts and approved the final manuscript.

## SUPPLEMENT—RESULT OF THE SEARCH

| Step | Searches | Results |
| --- | --- | --- |
| 1 | pandemic control.mp. [mp=ti, ab, hw, tn, ot, dm, mf, dv, kw, fx, dq, nm, kf, ox, px, rx, ui, sy] | 108 |
| 2 | pandemic interventions.mp. [mp=ti, ab, hw, tn, ot, dm, mf, dv, kw, fx, dq, nm, kf, ox, px, rx, ui, sy] | 15 |
| 3 | non-pharmaceutical interventions.mp. [mp=ti, ab, hw, tn, ot, dm, mf, dv, kw, fx, dq, nm, kf, ox, px, rx, ui, sy] | 283 |
| 4 | outbreak control.mp. [mp=ti, ab, hw, tn, ot, dm, mf, dv, kw, fx, dq, nm, kf, ox, px, rx, ui, sy] | 1314 |
| 5 | epidemic control.mp. [mp=ti, ab, hw, tn, ot, dm, mf, dv, kw, fx, dq, nm, kf, ox, px, rx, ui, sy] | 981 |
| 6 | epidemic interventions.mp. [mp=ti, ab, hw, tn, ot, dm, mf, dv, kw, fx, dq, nm, kf, ox, px, rx, ui, sy] | 30 |
| 7 | outbreak interventions.mp. [mp=ti, ab, hw, tn, ot, dm, mf, dv, kw, fx, dq, nm, kf, ox, px, rx, ui, sy] | 35 |
| 8 | 1 or 2 or 3 or 4 or 5 or 6 or 7 | 2742 |
| 9 | remove duplicates from 8 | 1653 |

## SUPPLEMENT—DESCRIPTION OF 3-STAGE FRAMEWORK

## REFERENCES

Armbruster B, Brandeau ML. Optimal mix of screening and contact tracing for endemic diseases. Math Biosci. 2007 Oct;209(2):386–402.

Barnes B, Glass K, Becker NG. The role of health care workers and antiviral drugs in the control of pandemic influenza. Math Biosci. 2007;209(2):403-416. doi:10.1016/j.mbs.2007.02.008

Becker NG, Glass K, Li Z, Aldis GK. Controlling emerging infectious diseases like SARS. Math Biosci. 2005;193(2):205–221. doi:10.1016/j.mbs.2004.07.006

Bell D, Nicoll A, et al. Non-pharmaceutical interventions for pandemic influenza, national and community measures. Emerg Infect Dis. 2006;12(1):88–94. doi:10.3201/eid1201.051371

Bell D, Nicoll A, et al. Non-pharmaceutical interventions for pandemic influenza, international measures. Emerg Infect Dis. 2006b;12(1):81-87. doi:10.3201/eid1201.051370

Bin Nafisah S, Alamery AH, Al Nafesa A, Aleid B, Brazanji NA. School closure during novel influenza: A systematic review. J Infect Public Health. 2018 Sep - Oct;11(5):657–661. doi: 10.1016/j.jiph.2018.01.003.

Bolton KJ, McCaw JM, Moss R, et al. Likely effectiveness of pharmaceutical and non-pharmaceutical interventions for mitigating influenza virus transmission in Mongolia. Bull World Health Organ. 2012;90(4):264–271. doi: 10.2471/BLT.11.093419

Bootsma MC, Ferguson NM. The effect of public health measures on the 1918 influenza pandemic in U.S. cities. Proc Natl Acad Sci USA. 2007;104(18):7588–7593. doi:10.1073/pnas.0611071104

Buonomo B, Messina E. Impact of vaccine arrival on the optimal control of a newly emerging infectious disease: A theoretical study. Math Biosci Eng. 2012;9(3):539–552. doi:10.3934/mbe.2012.9.539

Bwire G, Mwesawina M, Baluku Y, Kanyanda SS, Orach CG. Cross-Border Cholera Outbreaks in Sub-Saharan Africa, the Mystery behind the Silent Illness: What Needs to Be Done? PLoS One. 2016 Jun 3;11(6):e0156674. doi: 10.1371/journal.pone.0156674.

Caley P, Becker NG, Philp DJ. The waiting time for inter-country spread of pandemic influenza. PLoS One. 2007;2(1):e143. Published 2007 Jan 3. doi:10.1371/journal.pone.0000143

Chen T, Zhao B, Liu R, Zhang X, Xie Z, Chen S. Simulation of key interventions for seasonal influenza outbreak control at school in Changsha, China. J Int Med Res. 2020 Jan;48(1):300060518764268. doi: 10.1177/0300060518764268.

Chinazzi M, Davis JT, Ajelli M, Gioannini C, Litvinova M, Merler S, Pastore Y Piontti A, Mu K, Rossi L, Sun K, Viboud C, Xiong X, Yu H, Halloran ME, Longini IM Jr, Vespignani A. The effect of travel restrictions on the spread of the 2019 novel coronavirus (COVID-19) outbreak. Science. 2020 Mar 6. pii: eaba9757. Doi: 10.1126/science.aba9757.

Chong KC, Ying Zee BC. Modeling the impact of air, sea, and land travel restrictions supplemented by other interventions on the emergence of a new influenza pandemic virus. BMC Infect Dis. 2012 Nov 19;12:309. doi: 10.1186/1471-2334-12-309.

Dan YY, Tambyah PA, Sim J, Lim J, Hsu LY, Chow WL, Fisher DA, Wong YS, Ho KY. Cost-effectiveness analysis of hospital infection control response to an epidemic respiratory virus threat. Emerg Infect Dis. 2009 Dec;15(12):1909–16. doi: 10.3201/eid1512.090902.

Day T, Park A, Madras N, Gumel A, Wu J. When is quarantine a useful control strategy for emerging infectious diseases? Am J Epidemiol. 2006 Mar 1;163(5):479–85.

Figueroa A, Gulati RK, Rainey JJ. Estimating the frequency and characteristics of respiratory disease outbreaks at mass gatherings in the United States: Findings from a state and local health department assessment. PLoS One. 2017 Oct 27;12(10):e0186730. doi: 10.1371/journal.pone.0186730.

Gostic K, Gomez AC, Mummah RO, Kucharski AJ, Lloyd-Smith JO. Estimated effectiveness of symptom and risk screening to prevent the spread of COVID-19. Elife. 2020 Feb 24;9. pii: e55570. doi: 10.7554/eLife.55570.

Halder N, Kelso JK, Milne GJ. Cost-effective strategies for mitigating a future influenza pandemic with H1N1 2009 characteristics. PLoS One. 2011;6(7):e22087. doi: 10.1371/journal.pone.0022087.

Halton K, Sarna M, Barnett A, Leonardo L, Graves N. A systematic review of community-based interventions for emerging zoonotic infectious diseases in Southeast Asia. JBI Database System Rev Implement Rep. 2013 Feb;11(2):1–235. doi: 10.11124/jbisrir-2013-801. Epub 2013 Mar 12. PMCID: PMC4962925.

Handel A, Longini IM Jr, Antia R. What is the best control strategy for multiple infectious disease outbreaks? Proc Biol Sci. 2007 Mar 22;274(1611):833–7. doi: 10.1098/rspb.2006.0015.

Hellewell J, Abbott S, Gimma A, Bosse NI, Jarvis CI, Russell TW et al. Feasibility of controlling COVID-19 outbreaks by isolation of cases and contacts. The Lancet Global Health. 2020 Apr;pe488-e496. doi:10.1016/S2214-109X(20)30074-7

Herrera-Diestra JL, Meyers LA. Local risk perception enhances epidemic control. PLoS One. 2019 Dec 3;14(12):e0225576. doi: 10.1371/journal.pone.0225576.

Ishola DA, Phin N. Could influenza transmission be reduced by restricting mass gatherings? Towards an evidence-based policy framework. J Epidemiol Glob Health. 2011 Dec;1(1):33–60. doi: 10.1016/j.jegh.2011.06.004.

Jackson C, Mangtani P, Hawker J, Olowokure B, Vynnycky E. The effects of school closures on influenza outbreaks and pandemics: systematic review of simulation studies. PLoS One. 2014 May 15;9(5):e97297. doi: 10.1371/journal.pone.0097297.

Jefferson T, Del Mar CB, Dooley L, Ferroni E, Al-Ansary LA, Bawazeer GA, van Driel ML, Nair S, Jones MA, Thorning S, Conly JM. Physical interventions to interrupt or reduce the spread of respiratory viruses. Cochrane Database Syst Rev. 2011 Jul 6;(7):CD006207. doi: 10.1002/14651858.CD006207.pub4.

Khazeni N, Hutton DW, Garber AM, Hupert N, Owens DK. Effectiveness and cost-effectiveness of vaccination against pandemic influenza (H1N1) 2009. Ann Intern Med. 2009 Dec 15;151(12):829–39. Doi: 10.7326/0003-4819-151-12-200912150-00157.

Lee BY, Brown ST, Korch GW, Cooley PC, Zimmerman RK, Wheaton WD, Zimmer SM, Grefenstette JJ, Bailey RR, Assi TM, Burke DS. A computer simulation of vaccine prioritization, allocation, and rationing during the 2009 H1N1 influenza pandemic. Vaccine. 2010 Jul 12;28(31):4875–9. doi: 10.1016/j.vaccine.2010.05.002.

Lee BY, Wettstein ZS, McGlone SM, Bailey RR, Umscheid CA, Smith KJ, Muder RR. Economic value of norovirus outbreak control measures in healthcare settings. Clin Microbiol Infect. 2011 Apr;17(4):640–6. doi: 10.1111/j.1469-0691.2010.03345.x.

Lee VJ, Lye DC, Wilder-Smith A. Combination strategies for pandemic influenza response - a systematic review of mathematical modeling studies. BMC Med. 2009 Dec 10;7:76. doi: 10.1186/1741-7015-7-76.

Lempel H, Epstein JM, Hammond RA. Economic cost and health care workforce effects of school closures in the U.S. PLoS Curr. 2009 Oct 5;1:RRN1051.

Lin F, Muthuraman K, Lawley M. An optimal control theory approach to non-pharmaceutical interventions. BMC Infect Dis. 2010;10:32. Published 2010 Feb 19. doi:10.1186/1471-2334-10-32

MacIntyre CR, Chughtai AA. Facemasks for the prevention of infection in healthcare and community settings. BMJ. 2015 Apr 9;350:h694. doi: 10.1136/bmj.h694.

MacIntyre CR, Costantino V, Kunasekaran MP. Health system capacity in Sydney, Australia in the event of a biological attack with smallpox. PLoS One. 2019;14(6):e0217704. Published 2019 Jun 14. doi:10.1371/journal.pone.0217704

Mateus AL, Otete HE, Beck CR, Dolan GP, Nguyen-Van-Tam JS. Effectiveness of travel restrictions in the rapid containment of human influenza: a systematic review. Bull World Health Organ. 2014 Dec 1;92(12):868–880D. doi: 10.2471/BLT.14.135590.

Markel H, Lipman HB, Navarro JA, Sloan A, Michalsen JR, Stern AM, Cetron MS. Nonpharmaceutical interventions implemented by US cities during the 1918–1919 influenza pandemic. JAMA. 2007 Aug 8;298(6):644–54.

Madhav N, Oppenheim B, Gallivan M, Mulembakani P, Rubin E, Wolfe N. Chapter 17: Pandemics: Risks, Impacts, and Mitigation *in* Disease Control Priorities: Improving Health and Reducing Poverty. 3rd edition. Jamison DT, Gelband H, Horton S, et al., editors. Washington (DC): The International Bank for Reconstruction and Development / The World Bank; 2017 Nov 27.

Martinez DL, Das TK. Design of non-pharmaceutical intervention strategies for pandemic influenza outbreaks. BMC Public Health. 2014;14:1328. Published 2014 Dec 29. doi:10.1186/1471-2458-14-1328

Nguyen VK, Mikolajczyk R, Hernandez-Vargas EA. High-resolution epidemic simulation using within-host infection and contact data. BMC Public Health. 2018;18(1):886. Published 2018 Jul 17. doi:10.1186/s12889-018-5709-x

Moher D, Liberati A, Tetzlaff J, Altman DG; PRISMA Group. Preferred reporting items for systematic reviews and meta-analyses: the PRISMA statement. BMJ. 2009 Jul 21;339:b2535. doi: 10.1136/bmj.b2535.

Onder G, Rezza G, Brusaferro S. Case-Fatality Rate and Characteristics of Patients Dying in Relation to COVID-19 in Italy. JAMA. Published online March 23, 2020. doi: 10.1001/jama.2020.4683

Pan A, Liu L, Wang C, Guo H, Hao X, Wang Q, Huang J, He N, Yu H, Lin X, Wei S, Wu T. Association of Public Health Interventions With the Epidemiology of the COVID-19 Outbreak in Wuhan, China. JAMA. 2020 Apr 10. doi: 10.1001/jama.2020.6130.

Pasquini-Descomps H, Brender N, Maradan D. Value for Money in H1N1 Influenza: A Systematic Review of the Cost-Effectiveness of Pandemic Interventions. Value Health. 2017 Jun;20(6):819–827. doi: 10.1016/j.jval.2016.05.005.

Pérez Velasco R, Praditsitthikorn N, Wichmann K, Mohara A, Kotirum S, Tantivess S, Vallenas C, Harmanci H, Teerawattananon Y. Systematic review of economic evaluations of preparedness strategies and interventions against influenza pandemics. PLoS One. 2012;7(2):e30333. Doi: 10.1371/journal.pone.0030333.

Perlroth DJ, Glass RJ, Davey VJ, Cannon D, Garber AM, Owens DK. Health outcomes and costs of community mitigation strategies for an influenza pandemic in the United States. Clin Infect Dis. 2010 Jan 15;50(2):165–74. doi: 10.1086/649867.

Porgo TV, Norris SL, Salanti G, Johnson LF, Simpson JA, Low N, Egger M, Althaus CL. The use of mathematical modeling studies for evidence synthesis and guideline development: A glossary. Res Synth Methods. 2019 Mar;10(1):125–133. doi: 10.1002/jrsm.1333.

Prosser LA, Lavelle TA, Fiore AE, Bridges CB, Reed C, Jain S, Dunham KM, Meltzer MI. Cost-effectiveness of 2009 pandemic influenza A(H1N1) vaccination in the United States. PLoS One. 2011;6(7):e22308. doi: 10.1371/journal.pone.0022308.

Rainey JJ, Phelps T, Shi J. Mass Gatherings and Respiratory Disease Outbreaks in the United States - Should We Be Worried? Results from a Systematic Literature Review and Analysis of the National Outbreak Reporting System. PLoS One. 2016 Aug 18;11(8):e0160378. doi: 10.1371/journal.pone.0160378.

Rashid H, Ridda I, King C, Begun M, Tekin H, Wood JG, Booy R. Evidence compendium and advice on social distancing and other related measures for response to an influenza pandemic. Paediatr Respir Rev. 2015 Mar;16(2):119–26. doi: 10.1016/j.prrv.2014.01.003.

Ryu S, Gao H, Wong JY, Shiu EYC, Xiao J, Fong MW, et al. Nonpharmaceutical measures for pandemic influenza in nonhealthcare settings—international travel–related measures. Emerg Infect Dis. 2020 May. https://doi.org/10.3201/eid2605.190993

Sandford A. Coronavirus: Half of humanity now on lockdown as 90 countries call for confinement. Euronews. 2020 Apr 2. (https://www.euronews.com/2020/04/02/coronavirus-in-europe-spain-s-death-toll-hits-10-000-after-record-950-new-deaths-in-24-hou). Last accessed 2020 Jun 14.

Sacks JA, Zehe E, Redick C, Bah A, Cowger K, Camara M, Diallo A, Gigo AN, Dhillon RS, Liu A. Introduction of Mobile Health Tools to Support Ebola Surveillance and Contact Tracing in Guinea. Glob Health Sci Pract. 2015 Nov 12;3(4):646–59. doi: 10.9745/GHSP-D-15-00207.

Sang Z, Qiu Z, Yan X, Zou Y. Assessing the effect of non-pharmaceutical interventions on containing an emerging disease. Math Biosci Eng. 2012;9(1):147-164. doi:10.3934/mbe.2012.9.147

Saunders-Hastings P, Crispo JAG, Sikora L, Krewski D. Effectiveness of personal protective measures in reducing pandemic influenza transmission: A systematic review and meta-analysis. Epidemics. 2017 Sep;20:1–20. Doi: 10.1016/j.epidem.2017.04.003.

Saunders-Hastings P, Quinn Hayes B, Smith R, Krewski D. Modelling community-control strategies to protect hospital resources during an influenza pandemic in Ottawa, Canada. PLoS One. 2017;12(6):e0179315. Published 2017 Jun 14. doi:10.1371/journal.pone.0179315

Shi P, Keskinocak P, Swann JL, Lee BY. The impact of mass gatherings and holiday traveling on the course of an influenza pandemic: a computational model. BMC Public Health. 2010 Dec 21;10:778. doi: 10.1186/1471-2458-10-778.

Schiavo R, May Leung M, Brown M. Communicating risk and promoting disease mitigation measures in epidemics and emerging disease settings. Pathog Glob Health. 2014;108(2):76–94. doi:10.1179/2047773214Y.0000000127

Shiell A, Jorm LR, Carruthers R, Fitzsimmons GJ. Cost-effectiveness of measles outbreak intervention strategies. Aust N Z J Public Health. 1998;22(1):126-132. doi: 10.1111/j.1467-842x.1998.tb01156.x

Smith SM, Sonego S, Wallen GR, Waterer G, Cheng AC, Thompson P. Use of non-pharmaceutical interventions to reduce the transmission of influenza in adults: A systematic review. Respirology. 2015 Aug;20(6):896-903. doi: 10.1111/resp.12541.

Suphanchaimat R, Doung-Ngern P, Ploddi K, Suthachana S, Phaiyarom M, Pachanee K, Chaifoo W, Iamsirithaworn S. Cost effectiveness and budget impact analyses of influenza vaccination for prisoners in thailand: an application of system dynamic modelling. Int J Environ Res Public Health. 2020 Feb 14;17(4). pii: E1247. doi: 10.3390/ijerph17041247.

Townsend J, Greenland K, Curtis V. Costs of diarrhoea and acute respiratory infection attributable to not handwashing: the cases of India and China. Trop Med Int Health. 2017 Jan;22(1):74–81. doi: 10.1111/tmi.12808.

Tracht SM, Del Valle SY, Edwards BK. Economic analysis of the use of facemasks during pandemic (H1N1) 2009. J Theor Biol. 2012 May 7;300:161–72. doi: 10.1016/j.jtbi.2012.01.032.

Tuncer N, Mohanakumar C, Swanson S, Martcheva M. Efficacy of control measures in the control of Ebola, Liberia 2014–2015. J Biol Dyn. 2018;12(1):913–937. doi:10.1080/17513758.2018.1535095

van Genugten ML, Heijnen ML, Jager JC. Pandemic influenza and healthcare demand in the Netherlands: scenario analysis. Emerg Infect Dis. 2003;9(5):531-538. doi:10.3201/eid0905.020321

Viner RM, Russell SJ, Croker H, Packer J, Ward J, Stansfield C, Mytton O, Bonell C, Booy R. School closure and management practices during coronavirus outbreaks including COVID-19: a rapid systematic review. Lancet Child Adolesc Health. 2020 Apr 6. pii: S2352-4642(20)30095-X. Doi: 10.1016/S2352-4642(20)30095-X.

Wang B, Xie J, Fang P. Is a Mass Prevention and Control Program for Pandemic (H1N1) 2009 Good Value for Money? Evidence from the Chinese Experience. Iran J Public Health. 2012;41(11):34–43.

World Health Organization. Coronavirus disease 2019 (COVID-19) Situation Report – 77. 2020 Apr 6. (https://www.who.int/docs/default-source/coronaviruse/situation-reports/20200406-sitrep-77-covid-19.pdf). Last accessed 2020 Jun 14.

Young LS, Ruschel S, Yanchuk S, Pereira T. Consequences of delays and imperfect implementation of isolation in epidemic control. Sci Rep. 2019 Mar 5;9(1):3505. doi: 10.1038/s41598-019-39714-0.

Zhang Q, Wang D. Assessing the Role of Voluntary Self-Isolation in the Control of Pandemic Influenza Using a Household Epidemic Model. Int J Environ Res Public Health. 2015;12(8):9750–9767. Published 2015 Aug 18. doi:10.3390/ijerph120809750

Zhao S, Zhuang Z, Cao P, Ran J, Gao D, Lou Y, Yang L, Cai Y, Wang W, He D, Wang MH. Quantifying the association between domestic travel and the exportation of novel coronavirus (2019-nCoV) cases from Wuhan, China in 2020: a correlational analysis. J Travel Med. 2020 Mar 13;27(2). pii: taaa022. doi: 10.1093/jtm/taaa022.

